# Clinico-pathologic characteristics, patterns of treatment and outcome of newly diagnosed Waldenströms Macroglobulinemia- a single center real world retrospective analysis

**DOI:** 10.64898/2026.04.10.26350611

**Authors:** Vaibhav Gupta, Dibakar Podder, Saswata Saha, Bikash Shah, Shouriyo Ghosh, Jeevan Kumar, Arjin Philips Jacoby, Arijit Nag, Debranjani Chattyopadhyay, Rizwan Javed, Ashish Rath, Subhosmito Chakraborty, Rakesh Demde, Sushant Vinarkar, Mayur Parihar, Lateef Zameer, Deepak Mishra, Mammen Chandy, Reena Nair

## Abstract

Waldenström macroglobulinemia (WM) is a rare indolent neoplasm characterized by presence of ≥ 10% lymphoid cells in BM that exhibit plasmacytoid or plasma cell differentiation that secretes an IgM monoclonal protein. This is a retrospective analysis of 89 patients of WM that describes the clinical and laboratory characteristics, treatment patterns and outcome of patients of WM. The median age of the entire cophort was 66 years with male predominance (67.4%). Most common presentations were symptoms pertaining to anemia (77.5%) and constitutional symptoms (33.7%). Median bone marrow lymphoplasmacytic cells were 41%. Positivity for MYD88 and CXCR4 mutations were seen in 81.8% and 2.4% cases. BR was the most common regimen used (52.8%). Overall response rates were seen at 87.8%. Median overall survival, progression free survival and time to next treatment is 8.49 years, 2.15 years and 3.88 years. BR regimen was associated with highest event free survival.

## Introduction

Waldenström macroglobulinemia (WM) is a rare neoplasm characterized lymphoplasmacytic lymphoma (LPL) that secretes an IgM monoclonal protein (1). The disease is named after Swedish physician Jan Gösta Waldenström, who described two patients, with oronasal bleeding (hyperviscosity), anemia, lymphadenopathy, hypofibrinogenemia, bone marrow (BM) lymphocytosis and elevated sedimentation rate in 1944 (2). The age adjusted incidence of WM 0.36/100,000 general population in the United States (3). The World Health Organisation (WHO) defines this entity as a LPL secretes IgM monoclonal protein of any size, presence of ≥ 10% lymphoid cells in BM that exhibit plasmacytoid or plasma cell differentiation and exclusion of other lymphoproliferative disease (1,4). Patients usually present in their 6th or 7th decade with symptoms related to infiltration of BM (anemia, leukopenia, thrombocytopenia) or reticuloendothelial system (lymphadenopathy and hepatosplenomegaly), B symptoms (fever, night sweats and significant weight loss), IgM related (hyperviscosity, cryoglobulinemia, neuropathy) (4). Patients who are asymptomatic but fulfills the laboratory criteria is known as smoldering macroglobulinemia (SM), whereas the term IgM monoclonal gammopathy of undetermined significance (MGUS) reserved for asymptomatic patients who has only IgM paraprotein with no features of BM infiltration (5).

Apart from the characteristic clinical presentation, the diagnosis is confirmed on a bone marrow or lymph node histology that shows infiltration by pleomorphic B cells at different stages of maturation. The cells express pan B cell markers (CD19, CD20), together with CD22+dim, CD25+, CD27+ and IgM+; other antigens such as FMC7, BCL2, PAX5, CD81 and CD79b are usually positive as well, while CD10, CD11c, CD103 and CD23 are mostly absent. There is also presence of clonally restricted plasma cells that express CD38, CD138 and low levels of CD19 and CD20. The plasma cell population characteristically lacks CD56 expression seen in plasma cell neoplasms (6). There are characteristic genetic alterations seen in patients of WM. A single nucleotide change from T to C in MYD88 gene located on chromosome 3p22.2 is seen in up to 93% patients whereas CXCR4 mutations are seen in 29% patients (7–9). MYD88 is a member of the toll-like receptor (TLR) pathway that triggers downstream signalling in the presence of pathogenic activation of TLR. Mutation in MYD88 leads to activation leads to auto-assembly of myddosome that trigger recruitment of bruton tyrosine kinase (BTK) and interleukin 1 receptor associate kinase (IRAK4/IRAK1) that leads to prosurvival signalling (10). Somatic mutations in CXCR4 occur in the C terminal region, which are similar to congenital WHIM syndrome. CXCR4 mutations lead to robust interaction between bone marrow stromal cells and WM cells resulting in higher disease burden and rare extramedullary involvement (11).

WM is an indolent disease and treated when there are specific indications (12,13). There is no standard of care in the treatment of WM. Traditionally rituximab monotherapy or combinations with alkylators or proteasome inhibitors were used for treatment with the majority of patients attaining partial response (14,15). Bruton tyrosine kinase inhibitors (BTKI) are a class of drugs that inhibits BTK that includes ibrutinib, acalabrutinib, zanubrutinib and pirtobrutinib (16). BTKI are found to have superior response rate and progression free survival (PFS) (17–20).

There are very limited studies from India reporting demographic data, clinical presentations, treatment patterns and outcomes of patients of WM (21–23). This is a retrospective audit from a tertiary care center from eastern India reporting the clinical presentation, treatment patterns and outcome of patients of WM.

## Materials and methods

### Study Population

We retrospectively reviewed the medical records of LPL from electronic medical records (EMR) from May 2011 to January 2026. Patients diagnosed as WM were included in this analysis. Patients of WM who came for second opinion, precursor conditions (smoldering macroglobulinemia and IgM MGUS) are excluded from the current analysis.

Relevant clinical, demographic and laboratory data were extracted from EMR. The variables included, age, gender, chief complaints, performance status, hemoglobin, total leukocyte count, absolute neutrophil and lymphocyte count, platelet count, presence or absence of abnormal cells in peripheral blood, cryoglobulins, calcium, creatinine, beta2 microglobulin, immunoglobulin (Ig) G, A, M, serum protein electrophoresis, serum free light chain assay, lactate dehydrogenase, lymphoplasmacytoid (LP) cells on bone marrow aspiration and trephine biopsy, LP cells clonality assessment by immunohistochemistry (IHC) or flow cytometry, assessment of mutation for MYD88 and CXCR4 by allele specific polymerase chain reaction and Sanger sequencing respectively, histopathological examination and IHC on biopsy block, computed tomography (CT) or fluorodeoxyglucose positron emission tomography FDG PET CT finding, risk stratification as per International prognostic scoring system for Waldenström macroglobulinemia (IPSS-WM) and Modified Staging System for WM (MSS-WM), treatment received and response rates as per International Workshop on Waldenstrom’s Macroglobulinemia 2013 (IWWG) (24–26). Prior to data collection, approval from the institutional review board (IRB) was taken. Patient confidentiality and privacy were protected through de-identification of the data from the EMR. The study conforms to the declarations of Helsinki(27).

### Statistical analysis

Data were analyzed using SPSS v31 and R statistical software. Descriptive statistics and categorical variables were presented as frequencies and percentages. Survival outcomes including event free survival (EFS), overall survival (OS), time to next treatment (TTNT) estimated by kaplan meier method. EFS is calculated from the time of diagnosis to progression or death. OS was calculated from the date of diagnosis to the last follow up date or date of death. TTNT was calculated as the time interval between first line and second line treatment.

## Results

Baseline characteristics: From January 2011 to January 2026, we have diagnosed 123 cases of lymphoplasmacytic lymphoma, 108 patients (87%) were IgM secreting LPL. After excluding 4 cases of IgM MGUS and 15 cases of WM who came for second opinion, 89 cases of WM who were on regular follow up were included in the current analysis. The median age of the overall cohort was 66 years. Males were more commonly affected than females (M:F: 2.06:1). The most common presentation was with symptoms of anemia (77.5%%) followed by B symptoms (33.7%), symptoms of hyperviscosity (11.2%), bleeding manifestations (8.9%), neuropathy (8.9%), edema (5.6%), pleural/peritoneal effusion (3.3%), altered renal function, joint pain and leg ulcer were seen in 2.2% each. Lymphadenopathy, splenomegaly and hepatomegaly was seen in 33.7%%, 21.3% and 17.9% respectively. Table I

**Table I:** Baseline characteristics.

| Characteristics |  | n=89 (%) |
| --- | --- | --- |
| Age in years, Median (range) |  | 66 (45-83) |
| Gender | Male | 60 (67.4%) |
|  | Female | 29 (32.6%) |
| Clinical presentation | Symptomatic anemia | 69 (77.5%) |
|  | Lymphadenopathy | 30 (33.7%) |
|  | Constitutional symptoms | 30 (33.7%) |
|  | Splenomegaly | 19 (21.3%) |
|  | Hepatomegaly | 16 (17.9%) |
|  | Hyperviscosity | 10 (11.2%) |
|  | Bleeding | 8 (8.9%) |
|  | Neuropathy | 8 (8.9%) |
|  | Edema | 5 (5.6%) |
|  | Pleural effusion/ascites | 3 (3.3%) |
|  | Altered renal parameters | 2 (2.2%) |
|  | Leg ulcer | 2 (2.2%) |
|  | Joint pain | 2 (2.2%) |
| Laboratory characteristics |  |  |
| Hemoglobin (g/dL) median, range |  | 8 (4.1-13.5) |
| Hemoglobin <11.5 g/dl |  | 87 (97.7%) |
| Platelet count <100000 cells per microL |  | 22 (22.5%) |
| Presence of abnormal cells in peripheral smear (N=85) |  | 49 (57.6%) |
| Calcium > 10.4 mg/dl (N=86) |  | 10 (13.9%) |
| Creatinine > 2 mg/dl |  | 8 (8.9%) |
| M band (g/dl), median (range) (N=71) |  | 2.4 (0.27-8.5) |
| IgM (g/dL), median, range (N=88) |  | 4.3 (0.3-13.2) |
| Light chain restriction (n=75) | kappa (%) | 60 (80%) |
|  | lambda (%) | 15 (20%) |
| Beta2 microglobulin (mg/L), median, range (N=64) |  | 4.66 (1.58-16.3) |
| Lactate dehydrogenase (U/L) (N=74) |  | 186 (88-1128) |
| Bone marrow involvement |  | 85 (95.5%) |
| Bone marrow lympho-plasmacytic cells (%), median, range |  | 41 (11-92) |
| Amyloid deposition |  | 3 (3.3%) |
| Cryoglobulin |  | 3 (3.3%) |
| Molecular genetics | MYD88 (N=77) | 63 (81.8%) |
|  | CXCR4 (N=41) | 1 (2.4%) |
| IPSS-WM (n=64) | Low | 4 (6.2%) |
|  | Intermediate | 13 (20.3%) |
|  | High | 47 (73.4%) |
| MSS-WM (n=72) | Low | 16 (22.2%) |
|  | Low-intermediate | 23 (31.9%) |
|  | Intermediate | 25 (34.7%) |
|  | High | 8 (11.1%) |
| IPSS-WM: International prognostic scoring system for Waldenström macroglobulinemia; MSS-WM: Modified Staging System for WM |  |  |

Laboratory characteristics: Median hemoglobin, total leukocyte count and platelet count were 8 gm/dL, 5,450 cells/microL and 168,000/microL respectively. Presence of abnormal lymphoid cells were seen in 57.6% of cases in peripheral smear examination. Renal dysfunction was present in 8.9% cases. Median creatinine was 0.92 mg/dL (range: 0.35-9.7 g/dL). Hypercalcemia was seen in 13.9% cases. The median M band was 2.4 gm/dL (0.27-8.5 gm/dL) whereas median IgM was 4.3 g/dL (range: 0.3-13.2 gm/dL). Bone marrow studies were done in 85 cases and in remaining cases peripheral blood flow cytometry was used to enumerate the lymphoplasmacytic cells. Median lymphoid cells constituted 41% of all nucleated cells (range: 11-92%). Three patients (3.3%) showed amyloid deposition on bone marrow. Three patients (3.3%) tested positive for cryoglobins. Light chain restriction was assessed by flow cytometry on bone marrow aspirate or by immunohistochemistry on trephine biopsy in 75 samples. Kappa restriction was more common than lambda and seen in 80% cases. Myd88 mutation was assessed by allele specific polymerase chain reaction (AS-PCR) in 77 patients, 63 patients (81.8%) tested positive for Myd88 L265P mutation. CXCR4 mutation was tested in 41 patients and one patient (32.4%) tested positive, remaining 40 cases were wild type. Risk stratification was done as per IPSS-WM and MSS-WM in 64 and 72 cases respectively. As per IPSS, 6.2%, 20.3% and 73.4% belonged to low, intermediate and high risk respectively, whereas as per MSS, 22.2%, 31.9%, 34.7% and 131.1% of patients were stratified as low, low-intermediate, intermediate and high risk respectively. Table I

Treatment patterns and response rates: A variety of regimens were used for treatment of WM. Most common regimen used were bendamustine-rituximab (BR) (52.8%) followed by dexamethasone-rituximab-cyclophosphamide (DRC) (15.7%), bortezomib-cyclophosphamide-dexamethasone (VCD) (12.3%), bortezomib-dexamethasone-rituximab (BDR) (12.3%), and Bruton tyrosine kinase inhibitor (BTKi) (6.7%). In all rituximab-containing regimen, rituximab was introduced after patients’ IgM level reduced to less than 4gm/dl, to prevent IgM flare. Response assessment was done in 82 patients and 6.1% patients achieved complete response (CR) whereas 23.2%, 50% and 8.5% patients achieved very good partial response (VGPR), partial response (PR) and minor response (MR) respectively. Stable disease and progressive disease was seen in 6.1% each cases respectively (Table II). In the BR cohort, response was evaluated in 44 patients and CR, VGPR, PR and MR are seen in 11.3%, 29.5%, 47.7% and 6.8% respectively. The response rates of the other regimens are highlighted in table III. In the entire cohort 39 patients (43.8%) required second line treatment. Two patients had high grade transformation to diffuse large B cell lymphoma (DLBCL).Twenty patients (22.5%) died in the overall cohort. Of these, eight deaths were attributable to progressive disease, six to treatment-related complications, and six to causes unrelated to Waldenström macroglobulinemia (WM) or its treatment.

**Table II:** Treatment, Response and Outcomes.

|  |  |  |
| --- | --- | --- |
| Treatment (N=89) | BR | 47 (52.8%) |
|  | DRC | 14 (15.7%) |
|  | VCD | 11 (12.3%) |
|  | BDR | 11 (12.3%) |
|  | BTKI | 6 (6.7%) |
| Plasma exchange |  | 6 (6.7%) |
| Response rates<br>(n=82) | Complete response (CR) | 5 (6.1%) |
|  | Very good partial response (VGPR) | 19 (23.2%) |
|  | Partial response (PR) | 41 (50%) |
|  | Minor response (MR) | 7 (8.5%) |
|  | Stable disease (SD) | 5 (6.1%) |
|  | Progressive disease (PD) | 5 (6.1%) |
|  | Major response (PR+VGPR+CR) | 65 (79.2%) |
|  | Overall response (MR+PR+VGPR+CR) | 72 (87.8%) |
| Second Line treatment |  | 39 (43.8%) |
| High grade transformation |  | 2 (2.2%) |
| Bendamustine-Rituximab (BR), Bortezomib-cyclophosphamide-dexamethasone (VCD), Dexamethasone-rituximab-cyclophosphamide (DRC), Bortezomib-dexamethasone-rituximab (BDR), Bruton tyrosine kinase inhibitor (BTKi), CR: complete response, VGPR: very good partial response, PR: partial response, MR: minor response, SD: stable disease, PD: progressive disease |  |  |

**Table III:** Response rates across regimens.

| Regimens<br>(n=82) | Response rates |  |  |  |  |  |
| --- | --- | --- | --- | --- | --- | --- |
|  | CR | VGPR | PR | MR | SD | PD |
| BR (n=44) | 5 (11.3%) | 13 (29.5%) | 21 (47.7%) | 3 (6.8%) | 1 (2.27%) | 1 (2.27%) |
| DRC (n=11) | - | - | 5 (45.4%) | 2 (18.2%) | 2 (18.2%) | 2 (18.2%) |
| VCD (n=11) | - | 2 (18.1%) | 8 (72.8%) | - | - | 1 (9.1%) |
| BDR (n=11) | - | 2 (18.2%) | 6 (54.4%) | 1 (9.1%) | 1 (9.1%) | 1 (9.1%) |
| BTKI (n=5) | - | 2 (40%) | 1 (20%) | 1 (20%) | 1 (20%) |  |

Survival analysis: With a median follow up of 3.75 years (95% CI: 2.26 – 5.24 years), median OS was 8.49 years (95% CI: 5.98 - 11), median EFS was 2.15 years (95% CI: 0.56-3.74). Median time to next treatment was 3.88 years (95% CI: 1.84-5.93). (Figure II) Median EFS of BR, VCD, DRC, BDR, BTKI is 6.17 years 2.09 years, 1.15 years, 1.87 years and 1.19 years respectively. (Table IV)

**Fig I:**
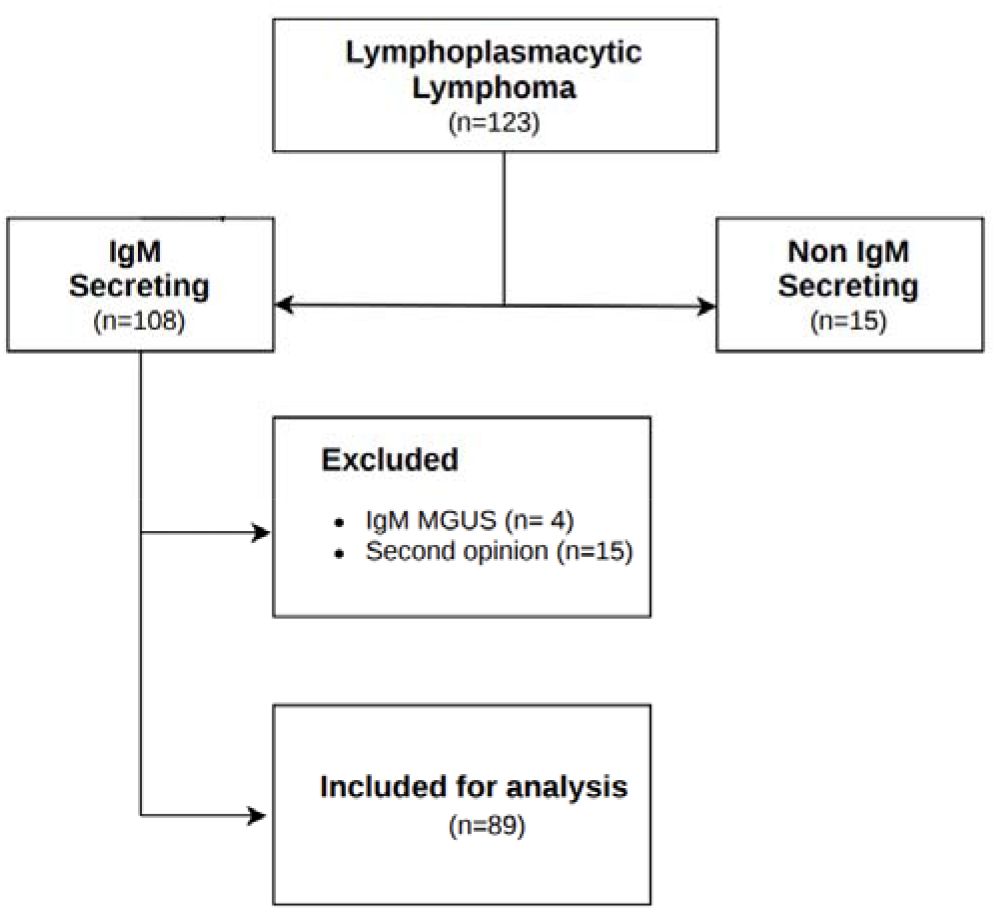
Consort diagram

**Fig II:**
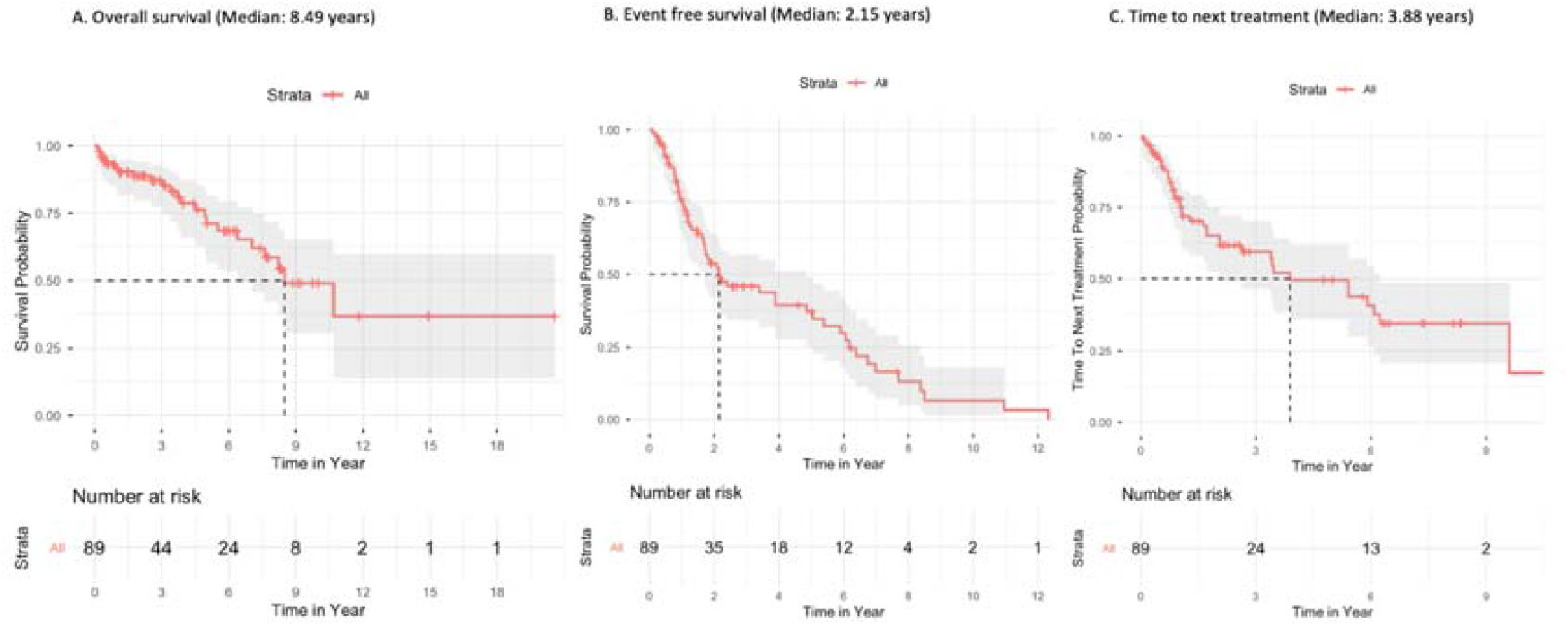
Overall survival, event free survival and time to next treatment

**Table IV:** Median event free survival (EFS) across regimens.

| Treatment | N | Median EFS (years) | 95% Confidence Interval |
| --- | --- | --- | --- |
| BR | 47 | 6.17 | 2.5-9.7 |
| VCD | 11 | 2.09 | 1.6-2.5 |
| DRC | 14 | 0.93 | 0.45-1.4 |
| BDR | 11 | 1.87 | 0.0-5.9 |
| BTKI | 6 | 0.63 | 0.0-1.54 |

## Discussion

The current study included 89 patients of WM where clinical, laboratory characteristics, patterns of treatment, responses and survival outcomes were analysed. The median age of 66 years and male preponderance correlates with other studies from India and registry data from Latin America and Europe (21–23,28,29). The most common clinical presentation in our cohort was symptomatic anemia (77.5%) followed by B symptoms (33.%), features of hyperviscosity (11.2%) and neuropathy (8.9%%). Lymphadenopathy, splenomegaly and hepatomegaly were seen in 33.7%, 21.63% and 17.9% respectively. These findings are also concordant with other studies by Prakash et al, Riva et al and Buske et al (22,30,31).

Almost all the patients (97.7%) had a hb less than 11.5 g/dl, thrombocytopenia was seen in 22.5% cases and presence of abnormal cells were seen in 57.6% cases. These are concordant with studies by Prakash et al and Riva et al (22,30). Hypercalcemia and renal dysfunction was seen in 13.9% and 8.9% cases. Although hypercalcemia and renal dysfunction is reported, the exact incidence is not known. In all the cases with hypercalcemia and renal dysfunction, plasma cell neoplasm is ruled out with confirmatory tests (immunohistochemistry and flow cytometry) and none of the cases had any bone lesion in imaging. We found a similar incidence of MYD88 mutation (81.8%) when compared to Treon et al, Riva et al and Patkar et al (7,23,30). We performed sanger sequencing for CXCR4 in 41 patients and only one patient was positive. This is in contrast to most studies where CXCR4 mutations are seen in almost 30% cases (5,11,32). In contrast to previous studies by Buske et al and Riva et al and Prakash et al, the current cohort had a higher proportion (73.4%) of high risk IPSSWM patients. This highlights higher disease bulk and probably late presentation to the clinic (22,30,31).

BR (52.8%) was the most common regimen used. In the overall cohort, major response (MaR) and overall response (OR) was seen in 79.2% and 87.8% respectively. This is similar to the study by Prakash et al where MaR and OR rates were 69.1% and 78.2% and they used BR in 61.1% of their patients (22). In the current study, 43.8% patients required second line treatment which is similar to the study by Prakash et al where 39.1% patients required second line therapy.

The median OS, EFS, TTNT in our cohort is 8.49 years, 2.15 years and 3.88 years. Although the OS and TTNT are comparable to other studies, EFS is variable across the literature. Studies by Buske et al, Kumar et al, reported similar PFS of 29 months and 22 months, whereas studies by Prakash et al and Riva et al reported a higher PFS of 45 months and 5 year PFS of 59% respectively (21,22,30,31). This higher variability may be due to patients’ risk at baseline and different regimens used for initial treatment.

To our knowledge, this analysis includes the highest number of patients of WM from India. But this study has limitations. Risk stratification was not uniformly done due to unavailability of beta2 microglobulin and lactate dehydrogenase, lack of MYD88 and CXCR4 testing and lack of response assessment in some patients.

## Conclusion

This study reiterates that there is less clinical heterogeneity in clinical findings when compared to published studies from high income countries. Bendamustine rituximab combinations produce deeper response and prolonged disease control when compared to other regimens. In the era of bruton tyrosine kinase inhibitors, the relevance of bendamustine-rituximab needs to be evaluated in clinical trials.

## Data Availability

All data produced in the present study are available upon reasonable request to the authors

## References

1. Owen RG, Treon SP, Al-Katib A, Fonseca R, Greipp PR, McMaster ML, et al. Clinicopathological definition of Waldenstrom’s macroglobulinemia: consensus panel recommendations from the Second International Workshop on Waldenstrom’s Macroglobulinemia. Semin Oncol. 2003 Apr;30(2):110–5. doi:10.1053/sonc.2003.50082 PubMed PMID: 12720118.

2. Waldenström J. Incipient myelomatosis or «essential«□hyperglobulinemia with fibrinogenopenia — a new syndrome? Acta Med Scand. 1944;117(3–4):216–47. doi:10.1111/j.0954-6820.1944.tb03955.x

3. McMaster ML. The epidemiology of Waldenström macroglobulinemia. Semin Hematol. 2023 Mar;60(2):65–72. doi:10.1053/j.seminhematol.2023.03.008

4. Kyle RA. Long-term natural history of Waldenstrom macroglobulinemia. Blood. 2003.

5. Gertz MA. Waldenström Macroglobulinemia: 2025 Update on Diagnosis, Risk Stratification, and Management. Am J Hematol. 2025;100(6):1061–73. doi:10.1002/ajh.27666

6. Dogliotti I, Jiménez C, Varettoni M, Talaulikar D, Bagratuni T, Ferrante M, et al. Diagnostics in Waldenström’s macroglobulinemia: a consensus statement of the European Consortium for Waldenström’s Macroglobulinemia. Leukemia. 2023 Feb;37(2):388–95. doi:10.1038/s41375-022-01762-3

7. Treon SP, Xu L, Yang G, Zhou Y, Liu X, Cao Y, et al. MYD88 L265P somatic mutation in Waldenström’s macroglobulinemia. N Engl J Med. 2012 Aug 30;367(9):826–33. doi:10.1056/NEJMoa1200710 PubMed PMID: 22931316.

8. Hunter ZR, Xu L, Yang G, Zhou Y, Liu X, Cao Y, et al. The genomic landscape of Waldenström macroglobulinemia is characterized by highly recurring MYD88 and WHIM-like CXCR4 mutations, and small somatic deletions associated with B-cell lymphomagenesis. Blood. 2014 Mar 13;123(11):1637–46. doi:10.1182/blood-2013-09-525808

9. Vinarkar S, Arora N, Chowdhury SS, Saha K, Pal B, Parihar M, et al. MYD88 and CXCR4 Mutation Profiling in Lymphoplasmacytic Lymphoma/Waldenstrom’s Macroglobulinaemia. Indian J Hematol Blood Transfus. 2019 Jan;35(1):57–65. doi:10.1007/s12288-018-0978-1 PubMed PMID: 30828149; PubMed Central PMCID: PMC6369099.

10. Treon SP, Cao Y, Xu L, Yang G, Liu X, Hunter ZR. Somatic mutations in MYD88 and CXCR4 are determinants of clinical presentation and overall survival in Waldenström macroglobulinemia. Blood. 2014 May 1;123(18):2791–6. doi:10.1182/blood-2014-01-550905

11. Treon SP, Gustine J, Meid K, Yang G, Xu L, Liu X, et al. Ibrutinib Monotherapy in Symptomatic, Treatment-Naïve Patients With Waldenström Macroglobulinemia. J Clin Oncol. 2018 Sep 20;36(27):2755–61. doi:10.1200/JCO.2018.78.6426

12. Castillo JJ, Advani RH, Branagan AR, Buske C, Dimopoulos MA, D’Sa S, et al. Consensus treatment recommendations from the tenth International Workshop for Waldenström Macroglobulinaemia. Lancet Haematol. 2020 Nov 1;7(11):e827–37. doi:10.1016/S2352-3026(20)30224-6 PubMed PMID: 33091356.

13. Pratt G, El-Sharkawi D, Kothari J, D’Sa S, Auer R, McCarthy H, et al. Diagnosis and management of Waldenström macroglobulinaemia-A British Society for Haematology guideline. Br J Haematol. 2022 Apr;197(2):171–87. doi:10.1111/bjh.18036 PubMed PMID: 35020191.

14. Paludo J, Abeykoon JP, Kumar S, Shreders A, Ailawadhi S, Gertz MA, et al. Dexamethasone, rituximab and cyclophosphamide for relapsed and/or refractory and treatment-naïve patients with Waldenstrom macroglobulinemia. Br J Haematol. 2017 Oct;179(1):98–105. doi:10.1111/bjh.14826 PubMed PMID: 28786474.

15. Buske C, Dimopoulos MA, Grunenberg A, Kastritis E, Tomowiak C, Mahé B, et al. Bortezomib-Dexamethasone, Rituximab, and Cyclophosphamide as First-Line Treatment for Waldenström’s Macroglobulinemia: A Prospectively Randomized Trial of the European Consortium for Waldenström’s Macroglobulinemia. J Clin Oncol. 2023 May 10;41(14):2607–16. doi:10.1200/JCO.22.01805

16. Castillo JJ, Buske C, Trotman J, Sarosiek S, Treon SP. Bruton tyrosine kinase inhibitors in the management of Waldenström macroglobulinemia. Am J Hematol. 2023 Feb;98(2):338–47. doi:10.1002/ajh.26788 PubMed PMID: 36415104; PubMed Central PMCID: PMC10107762.

17. Treon SP, Tripsas CK, Meid K, Warren D, Varma G, Green R, et al. Ibrutinib in Previously Treated Waldenström’s Macroglobulinemia. N Engl J Med. 2015 Apr 9;372(15):1430–40. doi:10.1056/NEJMoa1501548

18. Owen RG, McCarthy H, Rule S, D’Sa S, Thomas SK, Tournilhac O, et al. Acalabrutinib monotherapy in patients with Waldenström macroglobulinemia: a single-arm, multicentre, phase 2 study. Lancet Haematol. 2020 Feb 1;7(2):e112–21. doi:10.1016/S2352-3026(19)30210-8 PubMed PMID: 31866281.

19. Dimopoulos MA, Opat S, D’Sa S, Jurczak W, Lee HP, Cull G, et al. Zanubrutinib Versus Ibrutinib in Symptomatic Waldenström Macroglobulinemia: Final Analysis From the Randomized Phase III ASPEN Study. J Clin Oncol. 2023 Nov 20;41(33):5099–106. doi:10.1200/JCO.22.02830

20. Mato AR, Shah NN, Jurczak W, Cheah CY, Pagel JM, Woyach JA, et al. Pirtobrutinib in relapsed or refractory B-cell malignancies (BRUIN): a phase 1/2 study. Lancet Lond Engl. 2021 Mar 6;397(10277):892–901. doi:10.1016/S0140-6736(21)00224-5 PubMed PMID: 33676628; PubMed Central PMCID: PMC11758240.

21. Kumar S, Sanjeev, Rahman K, Singh MK, Chandra D, Gupta A, et al. Waldenström Macroglobulinemia: Clinico-pathological Profile and Treatment Outcomes of Patients from a Tertiary Care Centre of North India. Indian J Hematol Blood Transfus. 2021 Jul;37(3):386–90. doi:10.1007/s12288-020-01382-w PubMed PMID: 34267456; PubMed Central PMCID: PMC8239071.

22. Prakash G, Singh C, Reddy P, Lekshmon KS, Jain A, Khadwal A, et al. Clinical Characteristics and Treatment Outcomes of Patients with Waldenstrom Macroglobulinemia. Indian J Hematol Blood Transfus. 2025 Mar 29. doi:10.1007/s12288-025-02016-9

23. Patkar N, Subramanian PG, Deshpande P, Ghodke K, Tembhare P, Mascarenhas R, et al. MYD88 mutant lymphoplasmacytic lymphoma/Waldenström macroglobulinemia has distinct clinical and pathological features as compared to its mutation negative counterpart. Leuk Lymphoma. 2015 Feb 1;56(2):420–5. doi:10.3109/10428194.2014.924123 PubMed PMID: 24828863.

24. Owen RG, Kyle RA, Stone MJ, Rawstron AC, Leblond V, Merlini G, et al. Response assessment in Waldenström macroglobulinaemia: update from the VIth International Workshop. Br J Haematol. 2013;160(2):171–6. doi:10.1111/bjh.12102

25. Morel P, Duhamel A, Gobbi P, Dimopoulos MA, Dhodapkar MV, McCoy J, et al. International prognostic scoring system for Waldenstrom macroglobulinemia. Blood. 2009 Apr 30;113(18):4163–70. doi:10.1182/blood-2008-08-174961 PubMed PMID: 19196866.

26. Zanwar S, Le-Rademacher J, Durot E, D’Sa S, Abeykoon JP, Mondello P, et al. Simplified Risk Stratification Model for Patients With Waldenström Macroglobulinemia. J Clin Oncol. 2024 Jul 20;42(21):2527–36. doi:10.1200/JCO.23.02066

27. World Medical Association. World Medical Association Declaration of Helsinki: ethical principles for medical research involving human subjects. JAMA. 2013 Nov 27;310(20):2191–4. doi:10.1001/jama.2013.281053 PubMed PMID: 24141714.

28. Riva E. Treatment and survival outcomes of Waldenstrom macroglobulinemia in Latin America. JCO Glob Oncol. 2022.

29. Buske C, Sadullah S, Kastritis E, Tedeschi A, García-Sanz R, Bolkun L, et al. Treatment and outcome patterns in European patients with Waldenström’s macroglobulinaemia: a large, observational, retrospective chart review. Lancet Haematol. 2018 Jul 1;5(7):e299–309. doi:10.1016/S2352-3026(18)30087-5 PubMed PMID: 29958569.

30. Riva E, Duarte PJ, Valcárcel B, Remaggi G, Murrieta I, Corzo A, et al. Treatment and Survival Outcomes of Waldenstrom Macroglobulinemia in Latin American Patients: A Multinational Retrospective Cohort Study. JCO Glob Oncol. 2022 Aug 8;8:e2100380. doi:10.1200/GO.21.00380 PubMed PMID: 35939775; PubMed Central PMCID: PMC9470138.

31. Buske, Sadullah, Kastritis, Tedeschi, Et. A. Treatment and outcome patterns in European patients with Waldenström’s macroglobulinaemia: a large, observational, retrospective chart review. Lancet Haematol. 2018. doi:10.1016/s2352-3026(18)30087-5

32. Yan Y, Yu Y, Xiong W, Wang J, Yao Y, Jia Y, et al. Determination of MYD88 and CXCR4 Mutations for Clinical Detection and Their Significance in Waldenström Macroglobulinemia. Clin Cancer Res. 2024 Dec 2;30(23):5483–93. doi:10.1158/1078-0432.CCR-23-3939

